# Role of air temperature and humidity in the transmission of SARS-CoV-2 in the United States

**DOI:** 10.1101/2020.11.13.20231472

**Authors:** Yiqun Ma, Sen Pei, Jeffrey Shaman, Robert Dubrow, Kai Chen

**Affiliations:** Department of Environmental Health Sciences, Yale School of Public Health, 60 College Street, New Haven, CT, 06520-8034, USA; Yale Center on Climate Change and Health, Yale School of Public Health, 60 College Street, New Haven, CT, 06520-8034, USA; Department of Environmental Health Sciences, Mailman School of Public Health, Columbia University, New York, NY 10032, USA

## Abstract

Improved understanding of the effects of meteorological conditions on the transmission of SARS-CoV-2, the causative agent for COVID-19 disease, is urgently needed to inform mitigation efforts. Here, we estimated the relationship between air temperature or specific humidity (SH) and SARS-CoV-2 transmission in 913 U.S. counties with abundant reported infections from March 15 to August 31, 2020. Specifically, we quantified the associations of daily mean temperature and SH with daily estimates of the SARS-CoV-2 reproduction number (*Rt*) and calculated the fraction of *Rt* attributable to these meteorological conditions. Both lower temperature and lower SH were significantly associated with increased *Rt*. The fraction of *Rt* attributable to temperature was 5.10% (95% eCI: 5.00 - 5.18%), and the fraction of *Rt* attributable to SH was 14.47% (95% eCI: 14.37 - 14.54%). These fractions generally were higher in northern counties than in southern counties. Our findings indicate that cold and dry weather are moderately associated with increased SARS-CoV-2 transmissibility, with humidity playing a larger role than temperature.

## Introduction

Since emerging in Wuhan, China, the novel severe acute respiratory syndrome coronavirus 2 (SARS-CoV-2), the causative agent of coronavirus disease 2019 (COVID-19), has produced a major global pandemic. As of November 12, 2020, approximately 10.6 million COVID-19 cases and 243 thousand deaths had been reported in the U.S.^1^, more than any other country. The decreased stability of SARS-CoV-2 in warmer temperatures and higher humidity in laboratory experiments^2,3^, and the documented seasonality of influenza^4^ and infections caused by other coronaviruses^5-7^, lead to the hypothesis that lower air temperature and lower humidity are associated with increased SARS-CoV-2 transmission. Quantifying this effect on a population level is urgently needed to help inform public health control efforts, including transmission prevention and communication with the public^8^.

Numerous preliminary studies have found either positive or negative associations of air temperature and humidity with COVID-19 cases^9-13^. However, given the large number of undocumented SARS-CoV-2 infections^14^, the variations in the lag between infection and symptom onset, and the inconsistent lag between testing and reporting, using daily new confirmed cases may not be optimal for examining meteorological effects^15^. As a result, a few studies have used the reproduction number to estimate SARS-CoV-2 transmissibility^16-18^. One study reported high daily air temperature and high daily relative humidity (RH) to be associated with a reduced daily effective reproduction number (the mean number of new infections caused by a single infected person in a population in which some individuals may no longer be susceptible due to acquired immunity^19^) for SARS-CoV-2 in both China and the U.S.^16^. However, two early studies focused on the first few of months of the pandemic found no association between temperature or humidity and the basic reproduction number (the mean number of new infections caused by a single infected person in a population in which everyone is assumed to be susceptible and no public health measures have been implemented)^17,18^.

Early analyses, in particular, should be interpreted with caution^8^, as the range of temperature and humidity measurements during the short observation period at the beginning of the pandemic was relatively narrow in most studies^9-12,16-18^, thus limiting the ability to detect associations between these meteorological variables and SARS-CoV-2 transmission. In addition, many previous studies (whether using COVID-19 cases or reproduction number as the outcome) controlled for no or only a few potential confounders^9-13,17,18^, which include other environmental factors, socioeconomic factors, temporal changes in population immunity, and implementation of public health interventions.

Furthermore, although most early studies found an association between air temperature or humidity and COVID-19 incidence, the fraction of cases or deaths attributable to meteorological conditions remains unclear. One modeling study predicted that as long as most of the population is susceptible to infection, any role of humidity in SARS-CoV-2 transmission would be overwhelmed by the lack of population immunity^20^. This prediction is supported by the rapid transmission of SARS-CoV-2 regardless of climate zone, including warmer locations such as tropical Brazil, India and southern states in the U.S. during the northern hemisphere summer^1^.

Here we investigate the association between air temperature or specific humidity (SH; the mass of water vapor in a unit mass of moist air [g/kg]) and SARS-CoV-2 transmission, as measured by the reproduction number *Rt* (the mean number of new infections caused by a single infected person, given the public health measures in place, in a population in which everyone is assumed to be susceptible). We estimate *Rt* in the 913 counties with at least 400 cumulative cases as of August 31, 2020 and calculate the fraction of *Rt* attributable to temperature or SH, adjusting for a wide range of potential confounders.

## Results

### Distribution of meteorological factors and Rt

From March 15 to August 31, 2020, a total of 4,903,520 cases of COVID-19 were reported in the 913 study counties (Extended Data Table 1). We estimated the county-specific *Rt* using a dynamic metapopulation model informed by human mobility data that represents the transmission of SARS-CoV-2 in the U.S. (see Methods). Mean daily *Rt* averaged over all counties and days during the study period was 1.40 and ranged from 0.46 to 5.43. Daily air temperature and SH also ranged widely (air temperature: - 14.61 - 39.98 °C; SH: 0.99 - 22.15 g/kg). Union County, New Jersey had the highest *Rt* averaged over the study period (Fig. 1a). The largest number of cumulative cases per 100,000 people was observed in Chattahoochee County, Georgia, while Taylor County, Florida had the lowest number (Fig. 1b). Southern counties generally were hotter and more humid than northern counties; whereas the western U.S., coastal counties generally were cooler and more humid than inland counties (Fig. 1c-d).

**Fig. 1.**
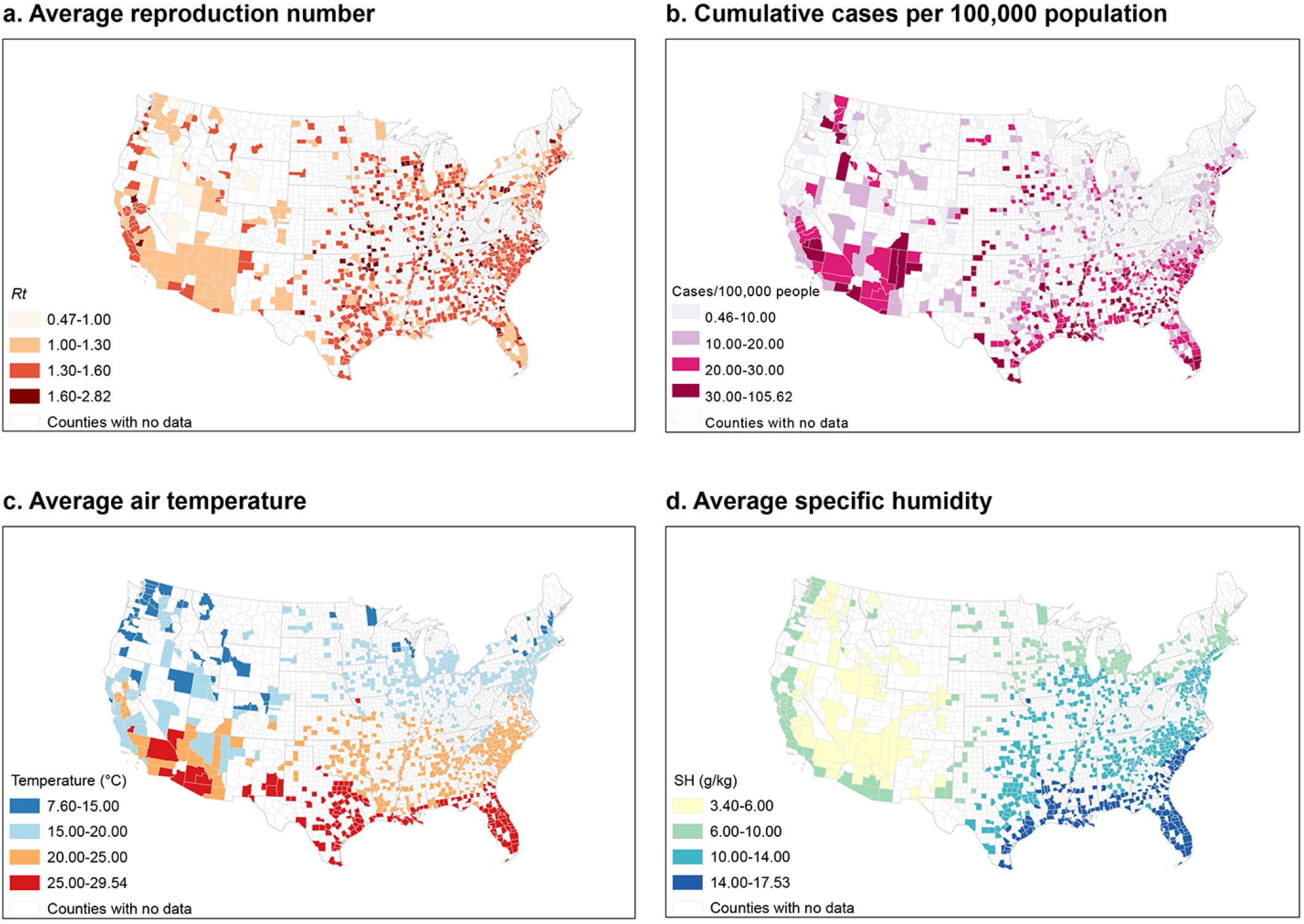
Map of the distribution of reproduction number, cumulative cases, air temperature and specific humidity in study counties. These maps display the distribution of the daily reproduction number (*Rt*), daily air temperature, and daily specific humidity (SH) averaged over the study period, and the cumulative cases per 100,000 population, in 913 U.S. counties.

### Associations between meteorological factors and Rt

We estimated the complex non-linear and temporally delayed associations of meteorological factors with the SARS-CoV-2 *Rt* using a generalized additive mixed model adjusting for spatiotemporal variations in *Rt* and potential measured confounders, described in detail in Methods. We then calculated the optimum values of temperature and SH, which correspond to the lowest *Rt*. We found an approximately linear inverse temperature-*Rt* relationship (Fig. 2a), with lower air temperatures significantly associated with increased transmission of SARS-CoV-2 when below the optimum temperature (32.57 °C). No significant associations were observed for temperatures above the optimum value. The relationship between SH and *Rt* was non-linear (Fig. 2b). Higher SH was significantly associated with decreased transmission, except for an increasing trend from approximately 9 to 15 g/kg. The optimum SH was estimated to be 19.78 g/kg. Compared with the optimum value, the 10^th^ percentile of the distribution of air temperature (8.8 °C) or SH (4.5 g/kg) was associated with a 14.10% (95% CI: 8.59 - 19.89%) and 27.49% (95% CI: 21.93 - 33.30%) increase of *Rt*, respectively. Effect estimates showed a decreasing trend in the lag dimension, diminishing to a small non-significant effect on lag day 13 (Extended Data Fig. 1). Sensitivity analyses showed the estimated relationships between air temperature or SH and *Rt* were generally consistent under different modeling choices (Fig. 2a-b).

**Fig. 2.**
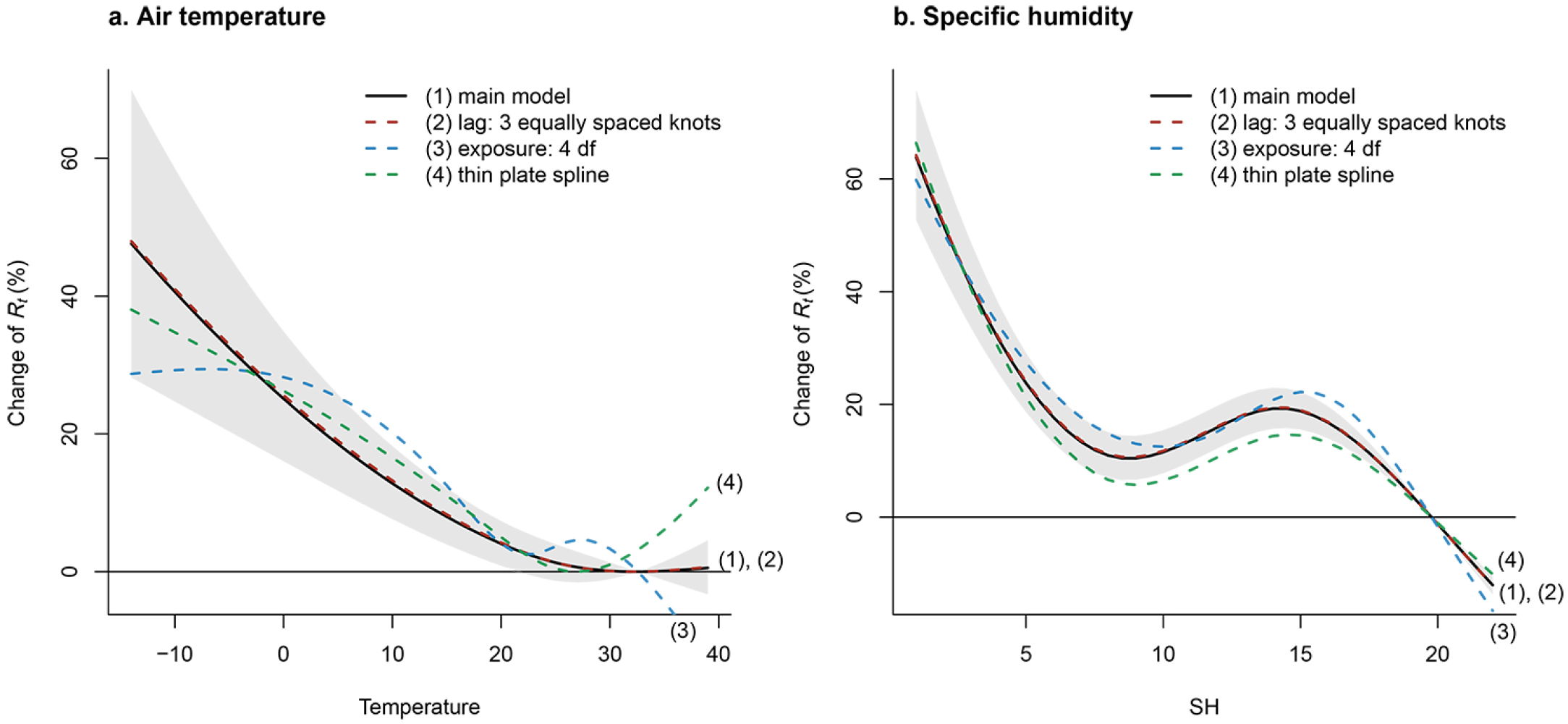
The associations of air temperature (°C) and specific humidity (g/kg) with *Rt*, under different choices of model. This figure shows the estimated exposure-response curves for the associations of air temperature (°C) and specific humidity (g/kg) with reproduction number (*Rt*) for SARS-Cov-2, with different modelling choices: (1) main model with 95% confidence interval (grey area): tensor product smooths to control for the temporal and spatial variations, and a cross-basis term for air temperature and SH, which is defined by natural cubic splines with 3 df for both the exposure-response and lag-response association, with a maximum lag of 13 days; (2) redefine the lag dimension using a natural cubic spline and 3 equally placed internal knots in the log scale; (3) change the df to 4 in the cross-basis term for air temperature or SH in the exposure-response function; (4): use a thin plate spline to control for geographical coordinates and time instead of using the tensor product smooths.

### Fractions of Rt attributable to meteorological factors

Based on the estimated exposure-response curves and daily county-specific *Rt*, we further calculated the fraction of *Rt* attributable to temperature or SH (i.e., the attributable fraction (AF), which can be interpreted as the fraction of *Rt* attributable to the deviation of temperature or SH from the optimum value). Across all 913 counties over the entire study period, the AF for temperature was 5.10% (95% empirical confidence intervals [95% eCI]: 5.00 - 5.18%), and the AF for SH was 14.47% (95% eCI: 14.37 - 14.54%) (Extended Data Table 2). The AF for temperature showed an increasing trend from south to north (Fig. 3a). The county with the highest AF for temperature was Whatcom County, Washington (12.88%) and the county with the lowest AF for temperature was Hidalgo County, Texas (0.52%). The AF for SH showed an increasing trend from south to north in the eastern U.S., whereas in the western U.S., the AF for SH was lower in counties in coastal states than in counties in interior states (Fig. 3b). The county with the highest AF for SH was Nye County, Nevada (27.47%), and the county with the lowest AF for SH was Plaquemines Parish, Louisiana (7.24%). The AF for temperature was the largest in March and April, and the lowest in July and August (Fig. 3c). The AF for SH showed a modest decline between March and August (Fig. 3d).

**Fig. 3.**
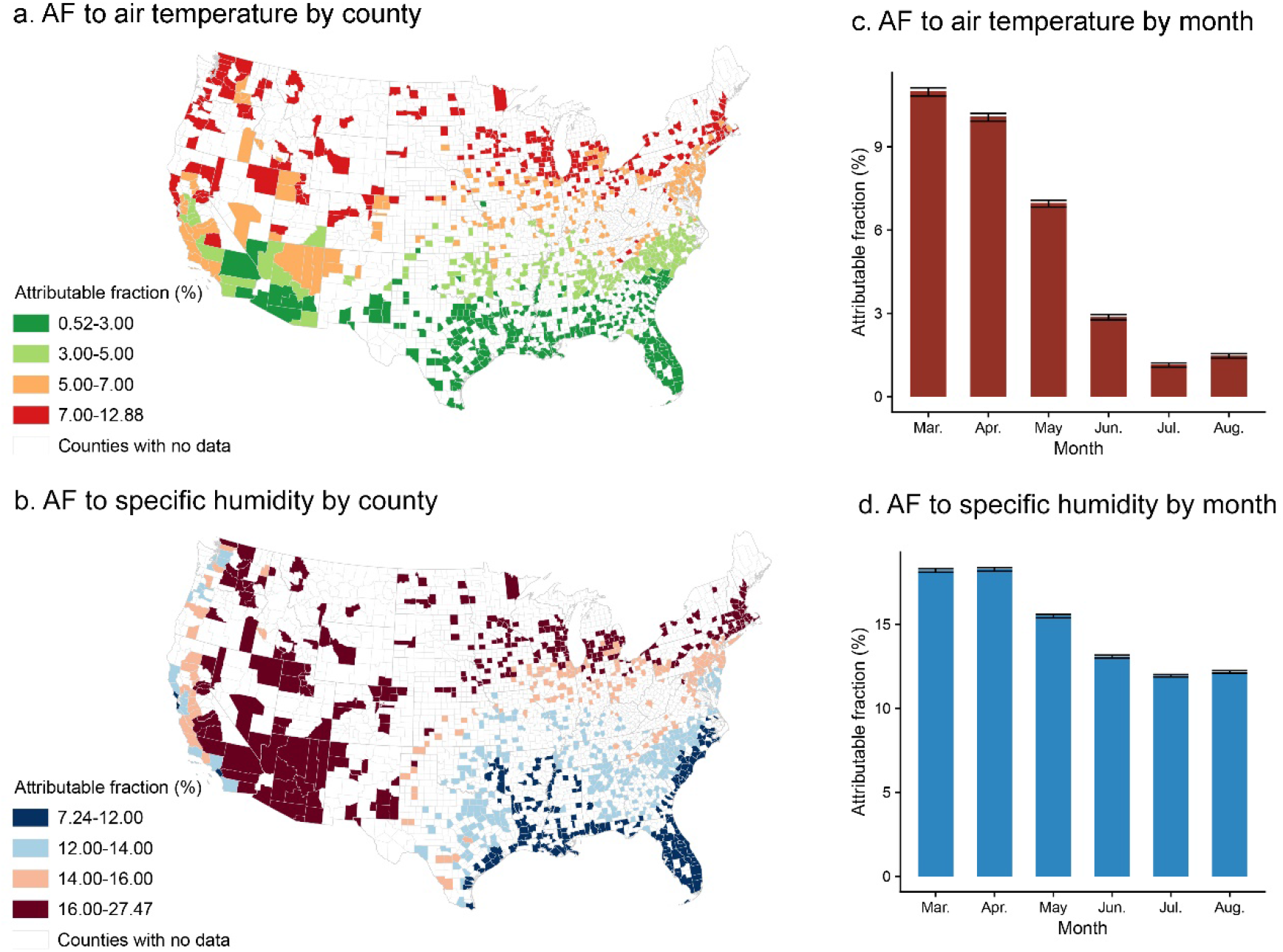
Fractions of *Rt* attributable to meteorological factors by county and month. a, b: the distribution of the fraction of reproduction number (*Rt*) attributable to temperature or specific humidity (i.e., attributable fraction [AF]) in each county; c, d: the distribution of AF across months in the study period. The black lines represent the 95% confidence interval, which were calculated by 1000 Monte Carlo simulations.

Sensitivity analyses indicate that the AF for air temperature remains robust when excluding socioeconomic factors and when additionally adjusting for smoking and obesity prevalence, long-term air pollution, or short-term air pollution (Extended Data Table 2). However, the estimated AF for temperature decreased from 5.11% to 3.55% after additionally adjusting for daily ultraviolet (UV). The AF for SH was robust across all sensitivity analyses.

## Discussion

Using estimated reproduction numbers for 913 U.S. counties and controlling for temporal and spatial trends and other potential confounders, we assessed the associations of air temperature and SH with the transmission of SARS-CoV-2 and estimated the fractions of *Rt* attributable to temperature and SH. We found both lower air temperature and lower SH to be significantly associated with increased *Rt*. During the study period, 5.10% of *Rt* was attributable to the deviation of temperature from its optimum value and 14.47% of *Rt* was attributable to the deviation of SH from its optimum value. Temperature and SH contributed more to transmission of SARS-CoV-2 in colder and drier counties and months than in warmer and more humid counties and months. In March (the coldest month of our study period of March-August), the AF for temperature was 11.00% and the AF for SH was 18.22% (Fig. 3). We can anticipate higher AFs during the colder and drier months of January and February.

Associations of lower temperature and lower humidity with increased COVID-19 outcomes have been reported by many previous studies. An early study in Wuhan, China reported that higher temperature and RH were associated with decreased COVID-19 deaths^21^. Many multicity analyses in China also supported such negative associations^9,11,12,22^. For example, using data of daily confirmed case counts from 30 provincial capital cities of China, Liu et al. found that lower temperature and lower absolute humidity were associated with higher COVID-19 case counts^11^. Later, with the rapid spread of COVID-19 around the world, studies in other countries emerged^23-25^. In the early stages of this pandemic in the U.S., a state-level study of daily COVID-19 case counts observed a declining trend of reported cases with increasing temperature up to 52 °F^23^. Based on data from 166 countries worldwide, another study reported that a 1 °C increase in temperature and a 1% increase in RH were associated with a 3.08% and 0.85% reduction in daily new cases, respectively^25^. However, many of these earlier studies were limited by short study periods (e.g. 1-2 months), use of daily confirmed cases or deaths across countries for which there were varying reporting biases, failure to account for the time lag between observed weather conditions and when cases or deaths were recorded, or failure to account for time delays between infection acquisition and case confirmation^15^.

By representing the transmissibility of SARS-CoV-2, the estimated daily reproduction number serves as a better outcome than daily case counts. While case counts are subject to the influence of reporting delay and underreporting, which vary across locations and are thus difficult to control, the reproduction number is a direct estimate of the transmission rate of SARS-CoV-2, quantifying the average number of infections caused by one infection in the population. A small number of studies previously analyzed the association between temperature or humidity and reproduction number^16-18^. Wang et al. found that a 1 °C increase in temperature was associated with a reduction in the effective reproduction number by 0.023 in China and 0.020 in the U.S., and a 1% increase in RH was associated with a reduction in the effective reproduction number by 0.0078 in China and 0.0080 in the U.S.^16^. These associations are consistent with our findings but were not supported by two studies in China that examined the basic reproduction number: the first found no association between temperature and SARS-CoV-2 transmission^18^; the second found no association between absolute humidity and SARS-CoV-2 transmission^17^. However, these early studies were limited by short observation periods at the beginning of the pandemic, and they did not account for variations of testing capacity, reporting, human mobility, and population susceptibility in estimating SARS-CoV-2 transmissibility.

In our study, *Rt* was estimated using a dynamic metapopulation model informed by human mobility data. This mechanistic model accounted for unreported infections, reporting delays, and county-to-county movement. We explicitly estimated the population susceptibility in each county, and removed its influence in the calculation of *Rt*^26^ (see Methods). Further, model estimated population susceptibility has been validated against independent seroprevalence studies^26^. Thus, our estimations account for spatial heterogeneity in population immunity.

Another strength of our study was adjustment for a wide range of demographic and socioeconomic factors in the main analysis, as well as for smoking and obesity, air pollution, and UV radiation in sensitivity analyses. We also thoroughly controlled for spatially and temporally heterogenous unmeasured confounders, such as implementation of and compliance with public health measures^26^, by simultaneously controlling for temporal and spatial variations and including a random intercept to further account for unmeasured county-level confounding (see Methods). This approach accounted for substantial differences in the epidemic curves among counties (see Extended Data Fig. 2).

Our findings are supported by laboratory evidence on the stability of SARS-CoV-2 as a function of temperature and humidity. It has been reported that the virus’ half-life in human nasal mucus and sputum is shorter under conditions of higher temperature and RH than under conditions of lower temperature and RH^2^. Similar findings were reported by other studies testing virus stability in virus transport medium^3^, in aerosols, and on various surfaces^27^. Further, the SARS-CoV-2 half-life was found to be longer at lower temperatures, and at both 22 °C and 27 °C, the half-life decreased as RH increased from 40% to 65% but increased as RH increased from 65% to 85%^28^. This result is consistent with the non-linear relationship between SH and *Rt* observed in our study (Fig. 2b), in which there was an increasing trend of *Rt* from 9 to 15 g/kg of SH superimposed on the overall decreasing trend.

The associations between temperature and humidity and SARS-CoV-2 transmissibility may be mediated by airway antiviral defenses. Inhalation of cold and dry air can impair mucociliary clearance, a crucial mechanism for elimination of inhaled pathogens^29^. Further, during the colder winter months people spend more time indoors, which may facilitate virus transmission^30^. During these months, whether indoors or outdoors, people are exposed to less UV radiation from the sun and therefore may produce less vitamin D and other UV-induced mediators of immune function^31^.

We found that SH contributes more to SARS-CoV-2 transmission than temperature, which is consistent with studies of influenza^32^. SH is more strongly associated with the observed seasonality of influenza in temperate regions than either temperature or RH^32-34^. In developed countries, such as the U.S., people spend approximately 90% of their time indoors^35^, especially during winter^30^. Although indoor temperature is usually controlled, indoor humidity generally is not, and closely mirrors outdoor levels^36-38^, perhaps explaining why ambient outdoor SH is more strongly associated with SARS-CoV-2 transmission than ambient outdoor temperature. However, it remains unclear whether SH is the causative modulator of SARS-CoV-2 transmission or is simply a useful indicator of the indoor environment and the combined effects of temperature and RH.

In the sensitivity analyses, after adjusting for daily UV radiation, the estimated AF for temperature decreased by about 30% (Extended Data Table 2), indicating that UV radiation acted as a confounder. This result is consistent with a recent study that found higher levels of solar UV radiation have a stronger association than temperature or humidity with a decreased growth rate (exponential increase in cases) of COVID-19^39^. In contrast, the fraction of *Rt* attributable to SH remained stable after adjusting for UV radiation. Although it is unclear why UV radiation would serve as a confounder for temperature, but not for SH, this result does suggest that SH is a more robust predictor than temperature.

Several limitations should be noted. First, this is an ecological rather than an individual-level study, thus making the study susceptible to the ecological fallacy. Second, due to the high correlation between temperature and SH, we were unable to explore whether the effects of temperature and humidity are independent. Third, our study period was restricted to March-August; if we had been able to include an entire year, including the colder months of November-February, our AF estimates for the entire study period would likely have been larger.

### Conclusion

Our findings indicate that cold and dry weather are moderately associated with increased SARS-CoV-2 transmissibility in the U.S., with absolute humidity (i.e., SH) playing a greater role than temperature. More extensive public health interventions are needed to mitigate the increased transmissibility of SARS-CoV-2 in winter months.

## Methods

### Data collection

We extracted hourly air temperature and SH from the North America Land Data Assimilation System project^40^, a near real-time dataset with a 0.125° × 0.125° grid resolution. We spatially and temporally averaged these data into daily county-level records. SH is the mass of water vapor in a unit mass of moist air (g/kg).

Other characteristics of each county, including geographic location, population density, demographic structure of the population, socioeconomic factors, intensive care unit (ICU) bed capacity, health risk factors, and long-term and short-term air pollution were collected from multiple sources. Geographic coordinates, population density, median household income, percent of people older than 60 years, percent black residents, percent Hispanic residents, percent owner-occupied housing, and percent residents aged 25 years and over without a high school diploma were collected from the U.S. Census Bureau^41^. The prevalence of smoking and obesity among adults in each county was obtained from the Robert Wood Johnson Foundation’s 2020 County Health Rankings^42^. Total ICU beds in each county were derived from Kaiser Health News^43^. We extracted annual PM_2.5_ concentrations in the U.S. from 2014 to 2018 from the 0.01° × 0.01° grid resolution PM_2.5_ estimation provided by the Atmospheric Composition Analysis Group^44^, and calculated average PM_2.5_ levels during this 5-year period for each county to represent long-term PM_2.5_ exposure (Extended Data Fig. 3). Short-term air quality data during the study period, including daily mean PM_2.5_ and daily maximum 8-hour O_3_, were obtained from the United States Environmental Protection Agency^45^. Daily downward UV radiation at the surface was extracted from the European Centre for Medium-Range Weather Forecasts ERA5 climate reanalysis^46^, with data available before August 2020.

### Estimation of reproduction number

We estimated the daily reproduction number (*Rt*) in all 3,142 U.S. counties using a dynamic metapopulation model informed by human mobility data^26^. *Rt* is the mean number of new infections caused by a single infected person, given the public health measures in place, in a population in which everyone is assumed to be susceptible. In the metapopulation model, two types of movement were considered: daily work commuting and random movement. The transmission dynamics are depicted by a set of ordinary differential equations^26^.

We explicitly simulated reported and unreported infections, for which separate transmission rates are defined, and allowed transmission rates and ascertainment rates to vary across different counties. To infer key epidemiological parameters, we fit the transmission model to county-level daily cases and deaths reported from March 15, 2020 to August 31, 2020. The estimated reproduction number is computed as *R*_*t*_ = *βD* [*α* +(1−*α*)*μ*], where *β* is county-specific transmission rate, *μ* is the relative transmissibility of unreported infections, *α* is the county-specific ascertainment rate, and *D* is the average duration of infectiousness. To avoid possible inaccurate estimation for counties with few cases, we inferred *Rt* in the 913 U.S. counties with at least 400 cumulative confirmed cases as of August 31, 2020 (Fig. 1). Details of the model fitting and *Rt* estimation are reported elsewhere^26^.

### Statistical analysis

All statistical analyses were conducted with R software (version 3.6.1) using the *mgcv* and *dlnm* package.

#### Exposure-response curves

Given the potential non-linear and temporally delayed effect of air temperature or SH, a distributed lag non-linear model (DLNM)^47^ combined with generalized additive mixed models (GAMM) was applied to estimate the associations of daily mean temperature or daily mean SH with SARS-CoV-2 *Rt*. Because of the high correlation between air temperature and SH (*r* = 0.80, Extended Data Table 3), we analyzed these two variables separately. The full model can be expressed as:

log(*E*(*R*_*i,t*_)) = *α* + *te*(*s*(latitude_*i*_, longitude_*i*_, k=30), *s*(time_*t*_, k=30)) + cb.temperature (or cb.SH) + *β*_1_(population density_*i*_) + *β*_2_(percent black residents_*i*_) + *β*_3_(percent Hispanic residents_*i*_) + *β*_4_(precent people older than 60 years_*i*_) + *β*_5_(median household income_*i*_) + *β*_6_(percent owner-occupied housing_*i*_) + *β*_7_(percent residents older than 25 years without a high school diploma_*i*_) + *β*_8_(number of ICU beds per 10,000 people_*i*_) + *u*_*i*_

where *E*(*R*_*i,t*_) refers to the expected *Rt* in county *i* on day *t*, and *α* is the intercept. The time trend was controlled by a flexible natural cubic spline over the range of study dates with a maximum of 30 knots; a thin plate spline with a maximum of 30 knots was used to control the coordinates of the centroid of each county. Due to the unique pattern of the non-linear time trend of *Rt* in each county (Extended Data Fig. 2), we constructed tensor product smooths (*te*) of the splines of geographical coordinates and time, to better control for the temporal and spatial variations. Cb.temperature or cb.SH is a cross-basis term for the mean air temperature or mean SH. We modeled exposure-response associations using a natural cubic spline with 3 degrees of freedom (df), and modeled the lag-response association using a natural cubic spline with an intercept and 3 df with a maximum lag of 13 days. We adjusted for county-level characteristics, including population density, percent black residents, percent Hispanic residents, percent people older than 60 years, median household income, percent owner-occupied housing, percent residents older than 25 years without a high school diploma, and number of ICU beds per 10,000 people. The random effect of county (*u*_*i*_) was considered in the model to further control for unmeasured county-level confounding. To obtain more precise estimates, we excluded from the analysis days during which *Rt* was less than 0.2.

Based on the estimated exposure-response curves, between the 1^st^ and the 99^th^ percentiles of the distribution of air temperature and SH, we determined the value of exposure associated with the lowest *Rt* to be the optimum temperature or the optimum SH, respectively. The natural cubic spline functions of the exposure-response relationship were then re-centered with the optimum temperature and SH as reference values. We report the cumulative relative risk of *Rt* associated with daily temperature or SH exposure in the previous two weeks (0 to 13 lag days) as the percent changes of *Rt* when comparing the daily exposure with the optimum reference values (i.e., the cumulative relative risk of *Rt* equals one and the percent change of *Rt* equals zero when the temperature or SH exposure is at its optimum value).

#### Attribution of Rt to temperature or SH

We used the optimum value of temperature or SH as the reference value for calculating the fraction of *Rt* attributable to temperature or SH; i.e., the attributable fraction (AF). For these calculations, we assumed that the associations of temperature and SH with *Rt* were consistent across the counties. For each day in each county, based on the cumulative lagged effect (cumulative relative risk) corresponding to the temperature or SH of that day, we calculated the attributable *Rt* in the current and next 13 days, using a previously established method^48^. Specifically, in a given county, the *Rt* attributable to a temperature or a SH (*x*_*t*_) for a given day *t* was defined as the attributable absolute excess of *Rt* (*AE*_*x,t*_, the excess reproduction number on day *t* attributable to the deviation of temperature or SH from the optimum value) and the attributable fraction of *Rt* (*AF*_*x*,_, the fraction of *Rt* attributable to the deviation of temperature or SH from the optimum value), each accumulated over the current and next 13 days. The formula can be expressed as

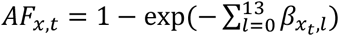 and 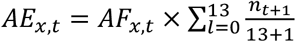, where *n*_*t*_ is the *Rt* on day *t*, and 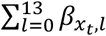 is the overall cumulative log-relative risk for exposure *x*_*t*_ on day *t* obtained by the exposure-response curves re-centered on the optimum values. Then, the total absolute excess of *Rt* attributable to temperature or SH in each county was calculated by summing the absolute excesses of all days during the study period, and the attributable fraction was calculated by dividing the total absolute excess of *Rt* for the county by the sum of the *Rt* of all days during the study period for the county. The attributable fraction for the 913 counties combined was calculated in a similar manner at the national level. We derived the 95% empirical confidence intervals (95% eCI) for the attributable absolute excess and attributable fraction by 1000 Monte Carlo simulations^48^. We also calculated the attributable fractions by month in the study period.

#### Sensitivity analyses

We conducted several sensitivity analyses to test the robustness of our results: a) the lag dimension was redefined using a natural cubic spline and three equally placed internal knots in the log scale; b) an alternative four df was used in the cross-basis term for air temperature or SH in the exposure-response function; c) time trend and geographical coordinates were controlled by a thin plate spline, instead of the tensor product smooth; d) all demographic and socioeconomic variables were excluded from the model; e) adjustment for the prevalence of smoking and obesity among adults was included in the model; f) additional adjustment was made for the average PM_2.5_ concentration in each county during 2014-2018^49^; g) additional adjustment was made for daily mean PM_2.5_, daily maximum 8-hour O_3_, and daily downward UV radiation at the surface. For daily covariates with available data in only some of the counties or study period, the results of sensitivity analyses were compared to the main model re-run on the same partial dataset.

## Data Availability

Estimates of county-level reproduction number are available at https://github.com/shaman-lab/Counterfactual
The data sets used in the study are publicly available from the following locations:
Hourly air temperature and specific humidity data: https://disc.gsfc.nasa.gov/datasets/NLDAS_FORA0125_H_002/summary?keywords=NLDAS
Population density, median household income, percent of people older than 60 years, percent black residents, percent Hispanic residents, percent owner-occupied housing, and percent residents aged 25 years and over without a high school diploma: https://www.census.gov/data/tables.html
U.S. county boundary: https://www.census.gov/geographies/mapping-files/time-series/geo/cartographic-boundary.html
Prevalence of smoking and obesity among adults in each county: https://www.countyhealthrankings.org/explore-health-rankings/rankings-data-documentation
Total ICU beds in each county: https://khn.org/news/as-coronavirus-spreads-widely-millions-of-older-americans-live-in-counties-with-no-icu-beds/
Annual PM2.5 concentrations in the U.S. from 2014 to 2018: http://fizz.phys.dal.ca/~atmos/martin/?page_id=140
Short-term daily mean PM2.5 and daily maximum 8-hour O3: https://www.epa.gov/outdoor-air-quality-data/download-daily-data
Daily downward UV radiation at the surface: https://cds.climate.copernicus.eu/cdsapp#!/dataset/reanalysis-era5-single-levels?tab=overview

## Acknowledgements

Y.M. received funding from the China Scholarship Council (201906320022). S.P. and J.S. acknowledged funding from the National Institutes of Health (GM110748) and the National Science Foundation (DMS-2027369), as well as a gift from the Morris-Singer Foundation. R.D. received funding from the High Tide Foundation.

## Author contributions

K.C. conceived of and supervised the conduct of this study and edited the manuscript. J.S. and R.D. contributed to the study design, interpretation of results, and manuscript revision. S.P. estimated the reproduction number and contributed to the writing. Y.M. conducted formal analyses and drafted the manuscript. All authors reviewed and approved the final version of this manuscript.

## Data availability

Estimates of county-level reproduction number are available at https://github.com/shaman-lab/Counterfactual

The data sets used in the study are publicly available from the following locations:

Hourly air temperature and specific humidity data: https://disc.gsfc.nasa.gov/datasets/NLDAS_FORA0125_H_002/summary?keywords=NLDAS

Population density, median household income, percent of people older than 60 years, percent black residents, percent Hispanic residents, percent owner-occupied housing, and percent residents aged 25 years and over without a high school diploma: https://www.census.gov/data/tables.html

U.S. county boundary: https://www.census.gov/geographies/mapping-files/time-series/geo/cartographic-boundary.html

Prevalence of smoking and obesity among adults in each county: https://www.countyhealthrankings.org/explore-health-rankings/rankings-data-documentation

Total ICU beds in each county: https://khn.org/news/as-coronavirus-spreads-widely-millions-of-older-americans-live-in-counties-with-no-icu-beds/

Annual PM_2.5_ concentrations in the U.S. from 2014 to 2018: http://fizz.phys.dal.ca/~atmos/martin/?page_id=140

Short-term daily mean PM_2.5_ and daily maximum 8-hour O_3_: https://www.epa.gov/outdoor-air-quality-data/download-daily-data

Daily downward UV radiation at the surface: https://cds.climate.copernicus.eu/cdsapp#!/dataset/reanalysis-era5-single-levels?tab=overview

## Code availability

R code for this analysis will be available at https://github.com/CHENlab-Yale/COVID-Climate

---

**Extended Data Fig. 1. Lag-response relationships of air temperature (°C) or specific humidity (g/kg) with reproduction number (*Rt*)**.

These curves are computed for the 10^th^ percentile of air temperature and specific humidity vs. the optimum values on different lag days; the grey areas display the 95% confidence interval. The effect estimates show a decreasing trend in the lag dimension, diminishing to a small non-significant effect on lag day 13.

**Extended Data Fig. 2. Daily *Rt* from March 15 to August 31 in the largest county in each state**.

Black dots represent the daily value of reproduction number (*Rt*) in the largest county in each U.S. state. Blue lines show the trend of *Rt* through time, fitted by local polynomial regression; the light blue areas display the 95% confidence interval.

**Extended Data Fig. 3. Distribution of average PM**_**2**.**5**_ **concentration during 2014-2018**.

This map displays the county-level average PM_2.5_ concentration during 2014-2018, extracted from the PM_2.5_ estimation provided by Atmospheric Composition Analysis Group.

