## Extended Data for "Role of air temperature and humidity in the transmission of SARS-CoV-2 in the United States"

**Extended Data Table 1. Summary of *Rt*, daily cases, meteorological variables, and other county-level characteristics in 913 U.S. counties**

| | Mean (SD) | Min | $P_{25}$ | Median | $P_{75}$ | Max |
|---|---|---|---|---|---|---|
| Daily records | | | | | | |
| *Rt* | 1.40 (0.55) | 0.46 | 1.00 | 1.36 | 1.74 | 5.43 |
| Cases (count) | 32.83 (126.50) | 0 | 1 | 6 | 22 | 9589 |
| Temperature (°C) | 20.48 (7.70) | -14.61 | 15.37 | 22.44 | 26.54 | 39.98 |
| SH (g/kg) | 11.18 (4.91) | 0.99 | 6.72 | 11.47 | 15.43 | 22.15 |
| County characteristics | | | | | | |
| Population density (people per square mile) | 556.16 (2649.60) | 1.95 | 68.74 | 161.82 | 424.00 | 72041.02 |
| Black residents (%) | 13.09 (14.72) | 0.12 | 2.49 | 7.33 | 18.96 | 78.51 |
| Hispanic residents (%) | 12.91 (15.01) | 0.24 | 3.89 | 6.99 | 15.74 | 95.49 |
| People aged 60+ (%) | 23.79 (5.06) | 7.09 | 20.58 | 23.49 | 26.24 | 50.22 |
| Median household income ($) | 559,29 (15094) | 277,95 | 45738 | 52,966 | 62,617 | 136,268 |
| Percent of owner-occupied housing (%) | 68.03 (7.76) | 22.77 | 63.61 | 68.81 | 73.42 | 85.34 |
| Residents over 25 years old without a high school diploma (%) | 12.81 (5.60) | 2.93 | 8.63 | 11.75 | 15.95 | 35.51 |
| ICU beds (per 10,000 people) | 0.23 (0.19) | 0.00 | 0.12 | 0.19 | 0.31 | 1.38 |

SD: standard deviation; Min: minimum; P25: 25th percentile; P75: 75th percentile; Max: maximum
SH: specific humidity
*Rt*: reproduction number

**Extended Data Table 2. Sensitivity analyses for attributable fraction of *Rt* across all 913 counties over the entire study period**

| | Air temperature | | | Specific humidity | | |
|---|---|---|---|---|---|---|
| | Optimum percentile | Optimum value (°C) | Total AF (%, 95% eCI) | Optimum percentile | Optimum value (g/kg) | Total AF (%, 95% eCI) |
| **Main model** | **99%** | **32.57** | **5.10 (5.00, 5.19)** | **99%** | **19.78** | **14.47 (14.37, 14.54)** |
| ***Adjust for or exclude county-level factors*[a]** | | | | | | |
| Exclude socioeconomic factors | 99% | 32.57 | 5.37 (5.26, 5.45) | 99% | 19.78 | 14.47 (14.37, 14.54) |
| Adjust for smoking and obesity prevalence | 99% | 32.57 | 5.13 (5.02, 5.21) | 99% | 19.78 | 14.49 (14.39, 14.57) |
| Adjust for long-term $PM_{2.5}$ | 99% | 32.57 | 5.40 (5.29, 5.48) | 99% | 19.78 | 14.54 (14.44, 14.61) |
| ***Adjust for daily UV radiation in days with available data*[b]** | | | | | | |
| Main model (UV) | 81% | 26.74 | 5.11 (5.03, 5.17) | 99% | 19.54 | 11.55 (11.44, 11.63) |
| Adjust for daily UV | 82% | 26.91 | 3.55 (3.47, 3.60) | 99% | 19.54 | 12.35 (12.24, 12.43) |
| ***Adjust for daily $PM_{2.5}$ in counties with available data*[c]** | | | | | | |
| Main model ($PM_{2.5}$) | 65% | 24.15 | 5.51 (5.42, 5.59) | 99% | 19.75 | 6.71 (6.47, 6.88) |
| Adjust for daily $PM_{2.5}$ | 65% | 24.15 | 5.52 (5.44, 5.59) | 99% | 19.75 | 6.79 (6.58, 6.95) |
| ***Adjust for daily $O_3$ in counties with available data*[d]** | | | | | | |
| Main model ($O_3$) | 54% | 22.63 | 3.61 (3.53, 3.67) | 99% | 19.81 | 7.03 (6.80, 7.20) |
| Adjust for daily $O_3$ | 54% | 22.63 | 3.59 (3.51, 3.65) | 99% | 19.81 | 7.03 (6.80, 7.19) |

*Rt*: reproduction number
AF: attributable fraction
eCI: empirical confidence interval
UV: ultraviolet
[a]: using full data with the same counties and period as in the main analysis.
[b]: using data in days with available daily UV radiation data (March 15 – August 31).
[c]: using data in counties with available daily $PM_{2.5}$ data (773 counties).
[d]: using data in counties with available daily $O_3$ data (771 counties).

**Extended Data Table 3a. Spearman correlation coefficients among main daily variables**

| | *Rt* | Daily cases | Temperature | SH |
|---|---|---|---|---|
| *Rt* | 1.00 | | | |
| Daily cases | -0.34 | 1.00 | | |
| Temperature | -0.19 | 0.27 | 1.00 | |
| SH | -0.10 | 0.21 | 0.80 | 1.00 |

SH: specific humidity
*Rt*: reproduction number

**Extended Data Table 3b. Spearman correlation coefficients among county-level mean *Rt* and other characteristics**

| | *Rt* | Population density | Black residents | Hispanic residents | People aged 60+ | Household income | House owner | Education | ICU beds |
|---|---|---|---|---|---|---|---|---|---|
| *Rt* | 1.00 | | | | | | | | |
| Population density | -0.01 | 1.00 | | | | | | | |
| Black residents | 0.01 | 0.27 | 1.00 | | | | | | |
| Hispanic residents | -0.27 | 0.08 | -0.20 | 1.00 | | | | | |
| People aged 60+ | 0.10 | -0.22 | -0.09 | -0.35 | 1.00 | | | | |
| Household income | -0.22 | 0.49 | -0.29 | 0.26 | -0.23 | 1.00 | | | |
| House owner | 0.02 | -0.17 | -0.23 | -0.21 | 0.46 | 0.16 | 1.00 | | |
| Education | 0.01 | -0.38 | 0.27 | 0.25 | -0.01 | -0.62 | -0.09 | 1.00 | |
| ICU beds | 0.09 | 0.21 | 0.24 | -0.12 | 0.03 | -0.24 | -0.36 | 0.01 | 1.00 |

*Rt*: reproduction number
ICU: intensive care unit

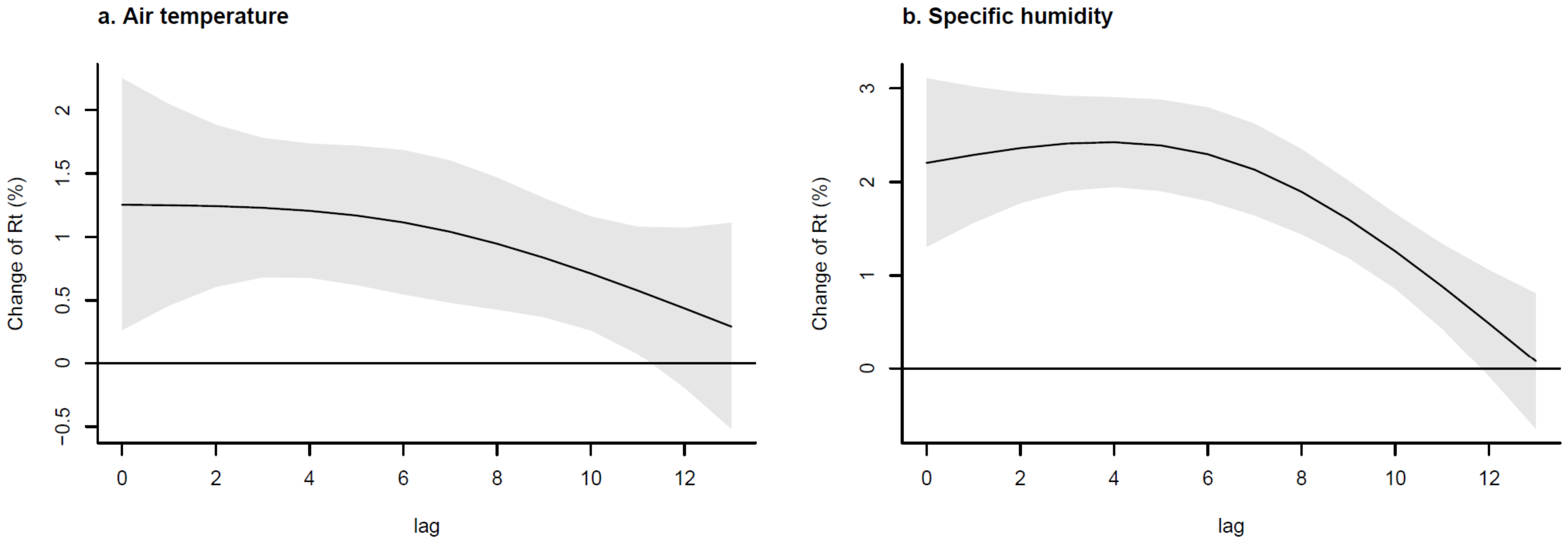

**Extended Data Fig. 1. Lag-response relationships of air temperature (°C) or specific humidity (g/kg) with reproduction number (*Rt*)**
These curves are computed for the 10th percentile of air temperature and specific humidity vs. the optimum values on different lag days; the grey areas display the 95% confidence interval. The effect estimates show a decreasing trend in the lag dimension, diminishing to a small non-significant effect on lag day 13.

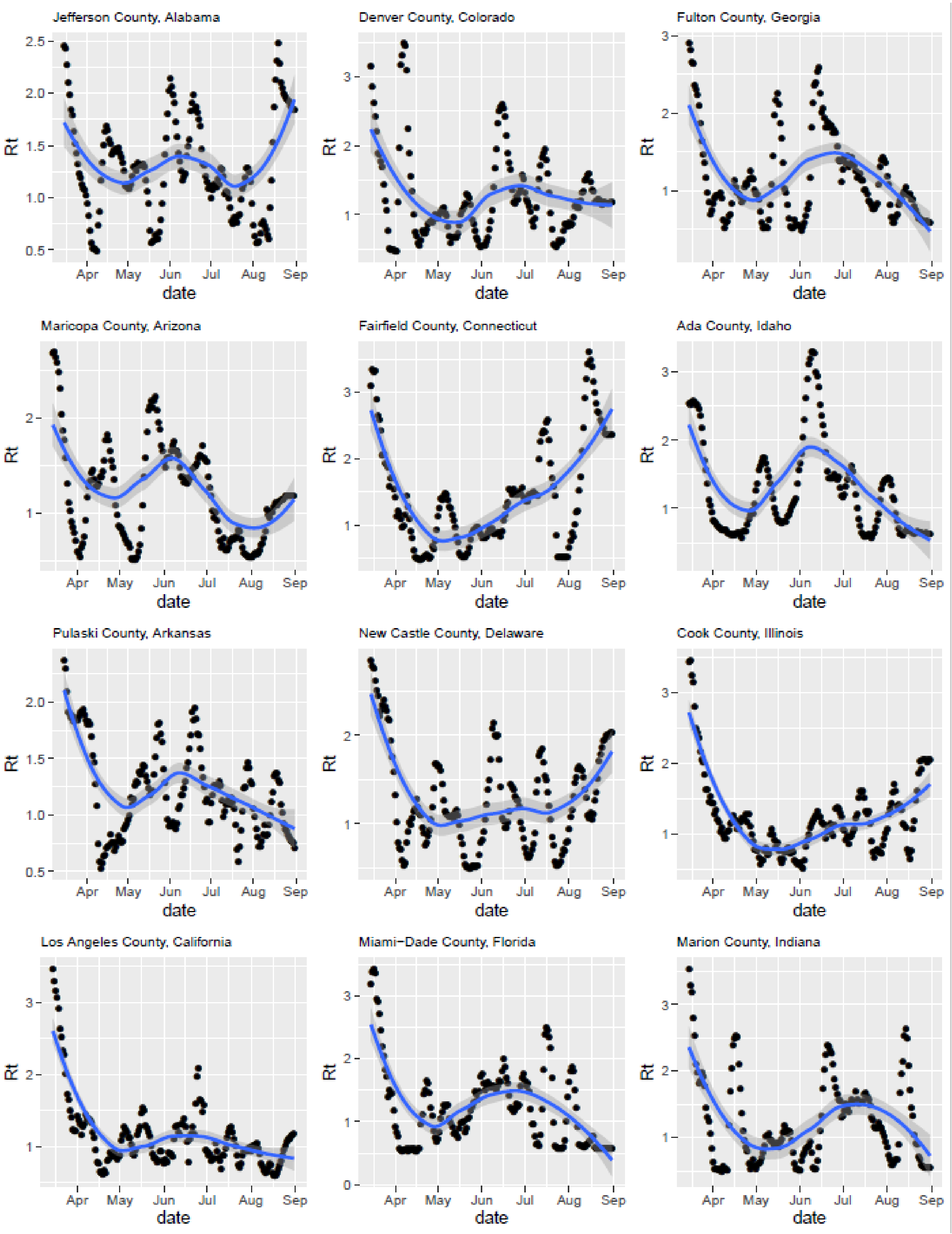
Jefferson County, Alabama
Denver County, Colorado
Fulton County, Georgia
Maricopa County, Arizona
Fairfield County, Connecticut
Ada County, Idaho
Pulaski County, Arkansas
New Castle County, Delaware
Cook County, Illinois
Los Angeles County, California
Miami−Dade County, Florida
Marion County, Indiana
Rt
date
Apr
May
Jun
Jul
Aug
Sep
0.5
1.0
1.5
2.0
2.5
0
1
2
3

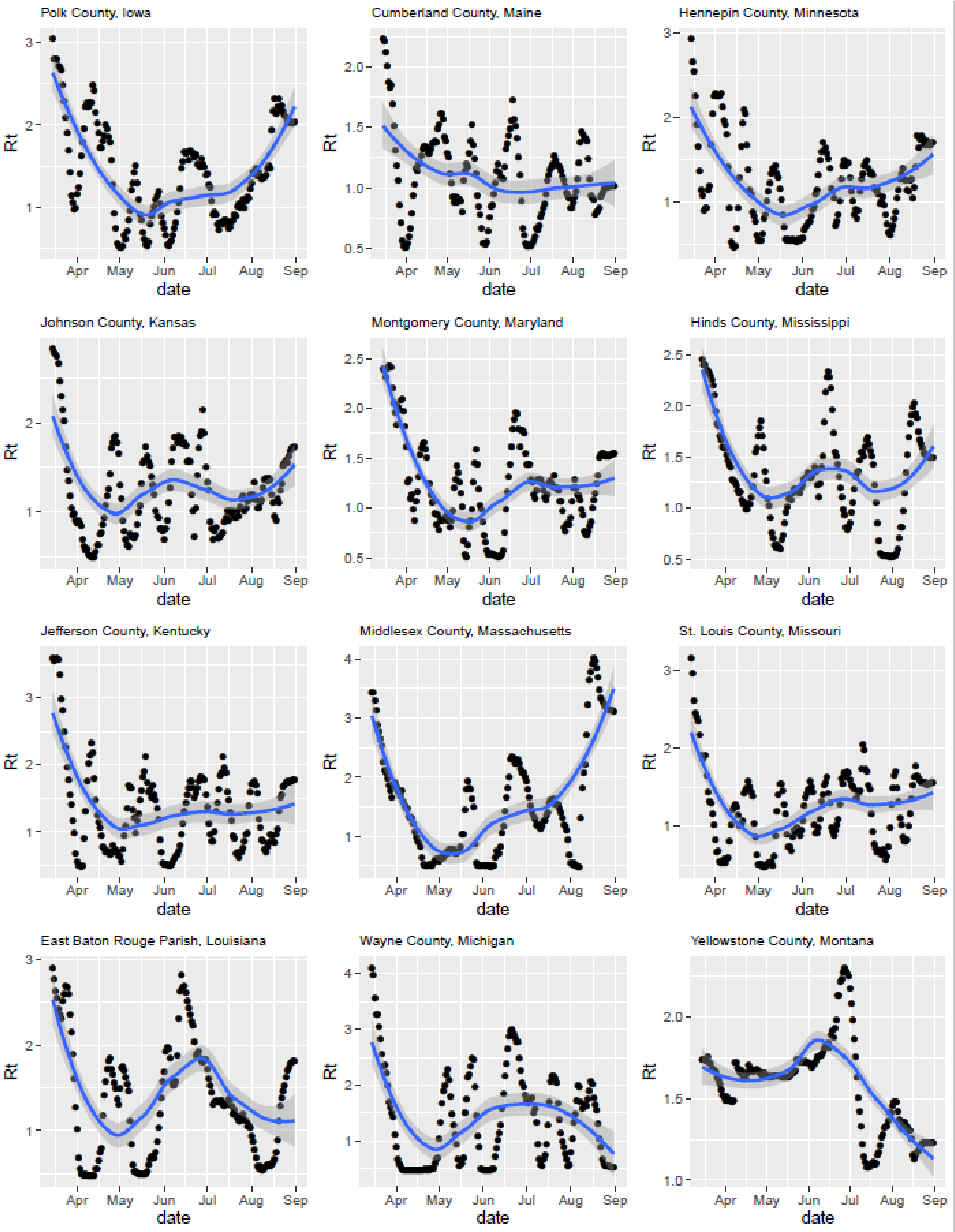
Polk County, Iowa
Cumberland County, Maine
Hennepin County, Minnesota
Johnson County, Kansas
Montgomery County, Maryland
Hinds County, Mississippi
Jefferson County, Kentucky
Middlesex County, Massachusetts
St. Louis County, Missouri
East Baton Rouge Parish, Louisiana
Wayne County, Michigan
Yellowstone County, Montana
Rt
date
Apr May Jun Jul Aug Sep

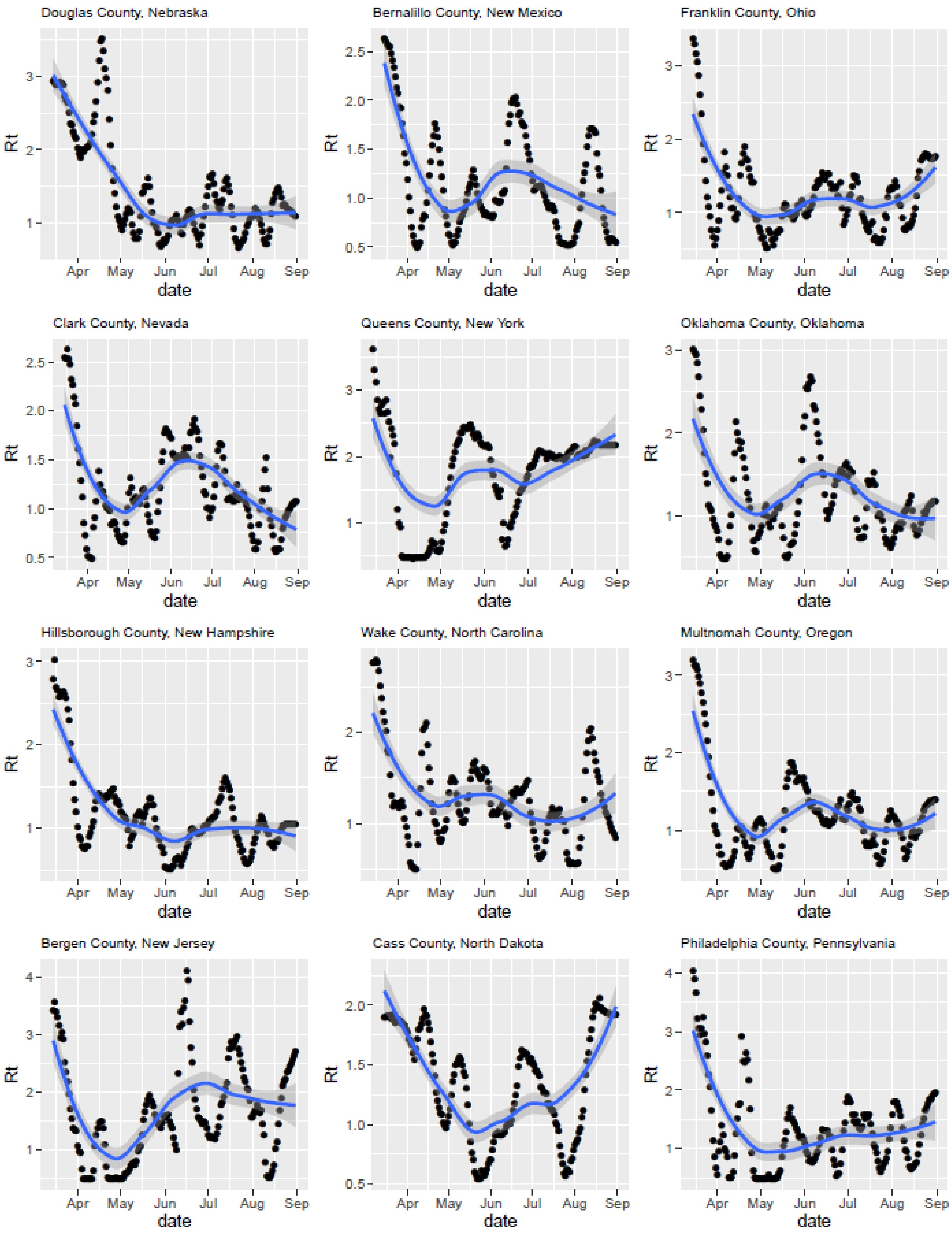
Douglas County, Nebraska
Bernalillo County, New Mexico
Franklin County, Ohio
Clark County, Nevada
Queens County, New York
Oklahoma County, Oklahoma
Hillsborough County, New Hampshire
Wake County, North Carolina
Multnomah County, Oregon
Bergen County, New Jersey
Cass County, North Dakota
Philadelphia County, Pennsylvania
Rt
date
Apr
May
Jun
Jul
Aug
Sep

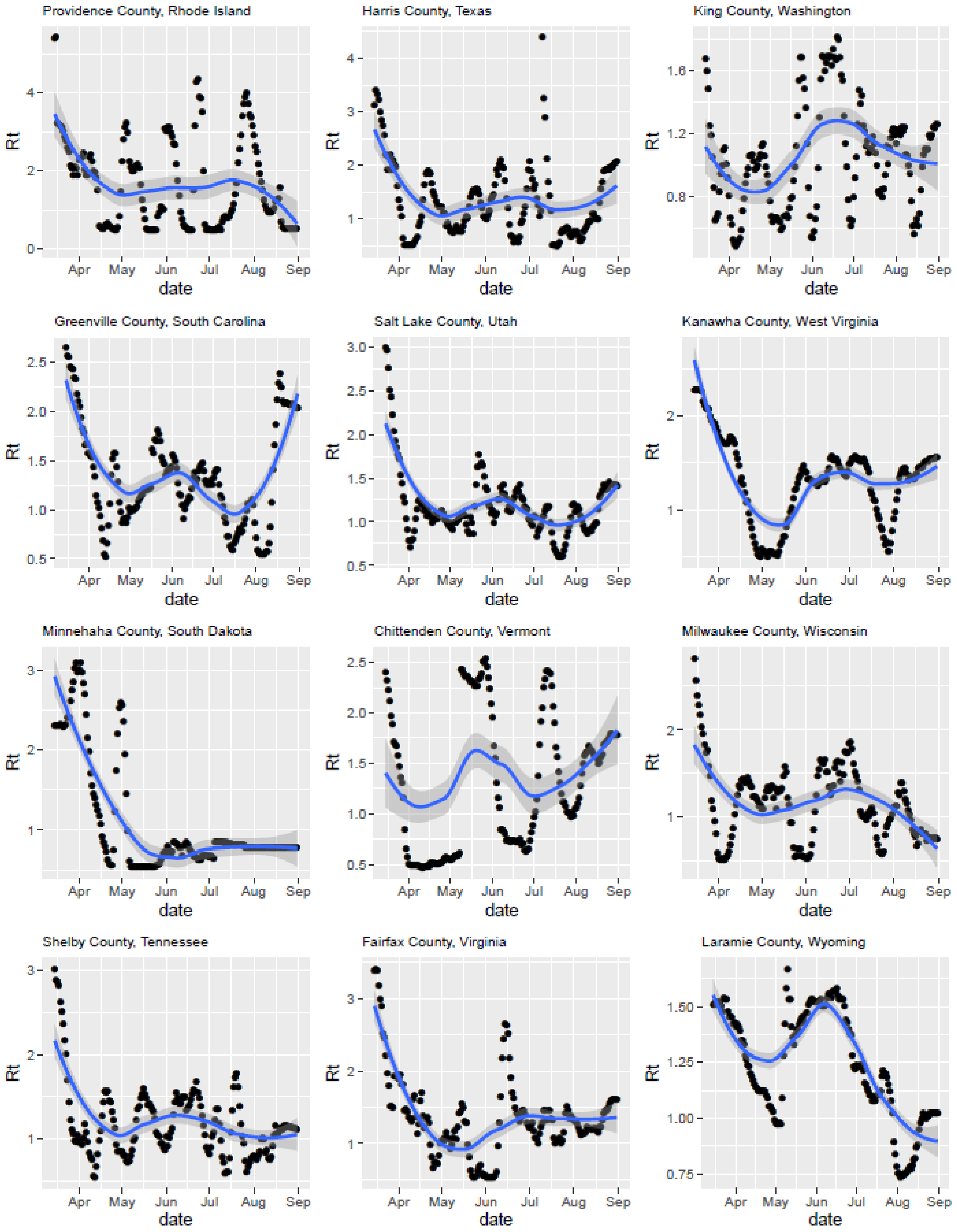

**Extended Data Fig. 2. Daily *Rt* from March 15 to August 31 in the largest county in each state**
Black dots represent the daily value of reproduction number (*Rt*) in the largest county in each U.S. state. Blue lines show the trend of *Rt* through time, fitted by local polynomial regression; the light blue areas display the 95% confidence interval.

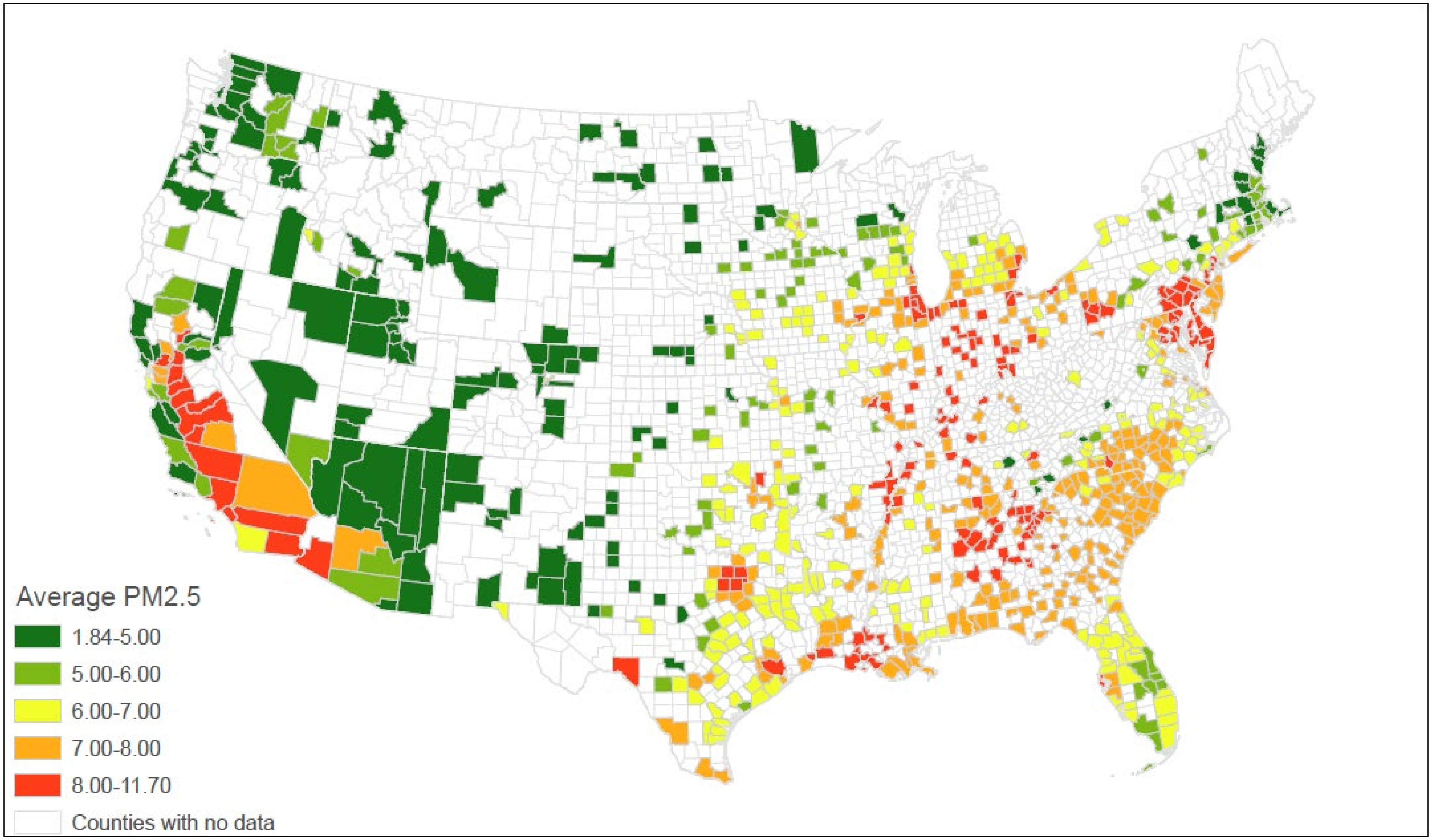

**Extended Data Fig. 3. Distribution of average $PM_{2.5}$ concentration during 2014-2018**

This map displays the county-level average $PM_{2.5}$ concentration during 2014-2018, extracted from the $PM_{2.5}$ estimation provided by Atmospheric Composition Analysis Group.
